# A Novel Fluorescent Immuno-Lectin Assay to Identify Osteoarthritis Associated Glycoforms of Lubricin

**DOI:** 10.1101/2024.01.02.23300279

**Authors:** Ali Reza Afshari, Kristina A. Thomsson, Jennifer Höglund, Henrik Ryberg, Kamlesh Gidwani, Kim Pettersson, Ola Rolfson, Lena I Björkman, Thomas Eisler, Tannin A. Schmidt, Gregory D. Jay, Niclas G. Karlsson

## Abstract

Lubricin or proteoglycan-4 (PRG-4) is a mucinous glycoprotein lubricating the cartilage and maintaining normal tissue function and cell-homeostasis. Altered *O*-glycoforms of lubricin has been found in osteoarthritis (OA) synovial fluid (SF). Here, we utilized a Fluorescent Immuno-Lectin Assay (FILA) to measure the levels of lubricin glycoforms in plasma/SF and their potential as biomarkers for disease. Five different lectins were used in the assay: Macrophage Galactose-type lectin (MGL), *Sambucus Nigra* Agglutinin (SNA), *Maackia Amurensis* Agglutinin (MAA), Peanut Agglutinin (PNA), and Galectin-3 (Gal-3). Our results showed that the levels of lubricin glycoforms in late-stage knee OA plasma (typically <10 μg/ml) are 1-3 order of magnitude lower than in OA SF (typically> 100 μg/ml). Furthermore, plasma lubricin glycans displayed higher level of sialylation, while SF derived lubricin displayed higher level of Tn-antigens. In OA patients we found decreased level in plasma of SNA binding lubricin (p=0.0023) compared to controls. This lectin binds 6 linked sialic acid. In addition, we found that lubricin glycoforms correlated both with Body Mass Index (BMI) and age, especially in regards of sialylation as measured by both MAA and SNA. Our data suggest that glycosylation of lubricin is different comparing SF with plasma. Moreover, the glycosylation of plasma lubricin is altered in OA patients compared controls.

## INTRODUCTION

Osteoarthritis (OA) is a degenerative joint disease where the protective cartilage that cushions the ends of the bones wears down and the underlying bone changes due to imbalance in cartilage catabolism^1,2^. The OA condition affects joints such as hands, knees, hips, and/or spine^1,3^. The seriousness of the disease is illustrated by that according to the World Health Organization (WHO) 9.6% of all 60+ years old men and 18% of all 60+ years old women are estimated to have symptomatic OA, worldwide^4^. The development of OA is believed to start as an initial alteration in pericellular matrix and softening of the extracellular matrix, with an increased mechanical stress of the chondrocyte and increased secretion of proteolytic enzyme degrading type II collagen and proteoglycans^5-7^. It is this process that eventually leads to a macroscopically detectable degradation of cartilage^8,9^. In addition to this pronounced degradation of the cartilage, OA also involves a low-grade joint inflammation, which manifests as an increased local level of inflammatory cytokines, chemokines, and growth factors^10^. At the present time, the diagnosis of OA is limited by radiographic evaluation, clinical examination, and detection of signs/symptoms such as crepitus, morning stiffness of joints, joint pain, joint instability, and bony enlargement (osteophytes)^3,11^. There are no approved molecular diagnostics for OA. In an early non-chronic stage of OA, there is a potential that diagnostics will be able to identify individuals that will benefit from future disease modifying osteoarthritis drugs (DMOADs) or can be selected to less invasive therapy compared to total joint replacement surgery^11,12^. Hence, there is a need to use modern type of omics methodology to identify biomarker candidates that could aid in identifying subgroups of patients that are at risk for developing OA and would benefit from precision medicine targeting their specific phenotype of the disease. We and others have pursued the use of glycomics for stratification of OA patients, specifically looking into the *O*-glycome of the lubricin in synovial fluid (SF)^13^. The SF provides the lubrication to the joint giving an almost friction free motion. There are mainly three types of biomolecules that have been implicated being involved in this lubrication: phospholipids^14^, hyaluronan (HA), and glycoproteins (lubricin^15^ and aggrecan^16^).

Lubricin (Figure 1) is the key *O*-glycoprotein responsible for boundary lubrication of diarthrodial joints and for the integrity of articular cartilage^15,17^. Lubricin has earned the attention for its involvement in joint degrading diseases such as OA and rheumatoid arthritis^18^. In the articular cartilage, lubricin is expressed by synoviocytes located on synovial membrane^19^ and chondrocytes embedded in cartilage matrix^20^. In addition to the joints, lubricin is widely distributed in the body, being expressed also by cells in the liver, heart, lungs, brain, prostate, small intestine, joints and tendon^21,22^. The expression of lubricin by various cells, alternative splicing, and post-translational modifications give rise to different isoforms of lubricin^23^. In accordance with lubricin’s wide distribution, the protein has been implicated in other non-joint associate diseases, where its regulation in plasma and liver in a murine sepsis model is in line with the role of lubricin as an immune- and inflammatory-regulator^24,25^.

**Figure 1:**
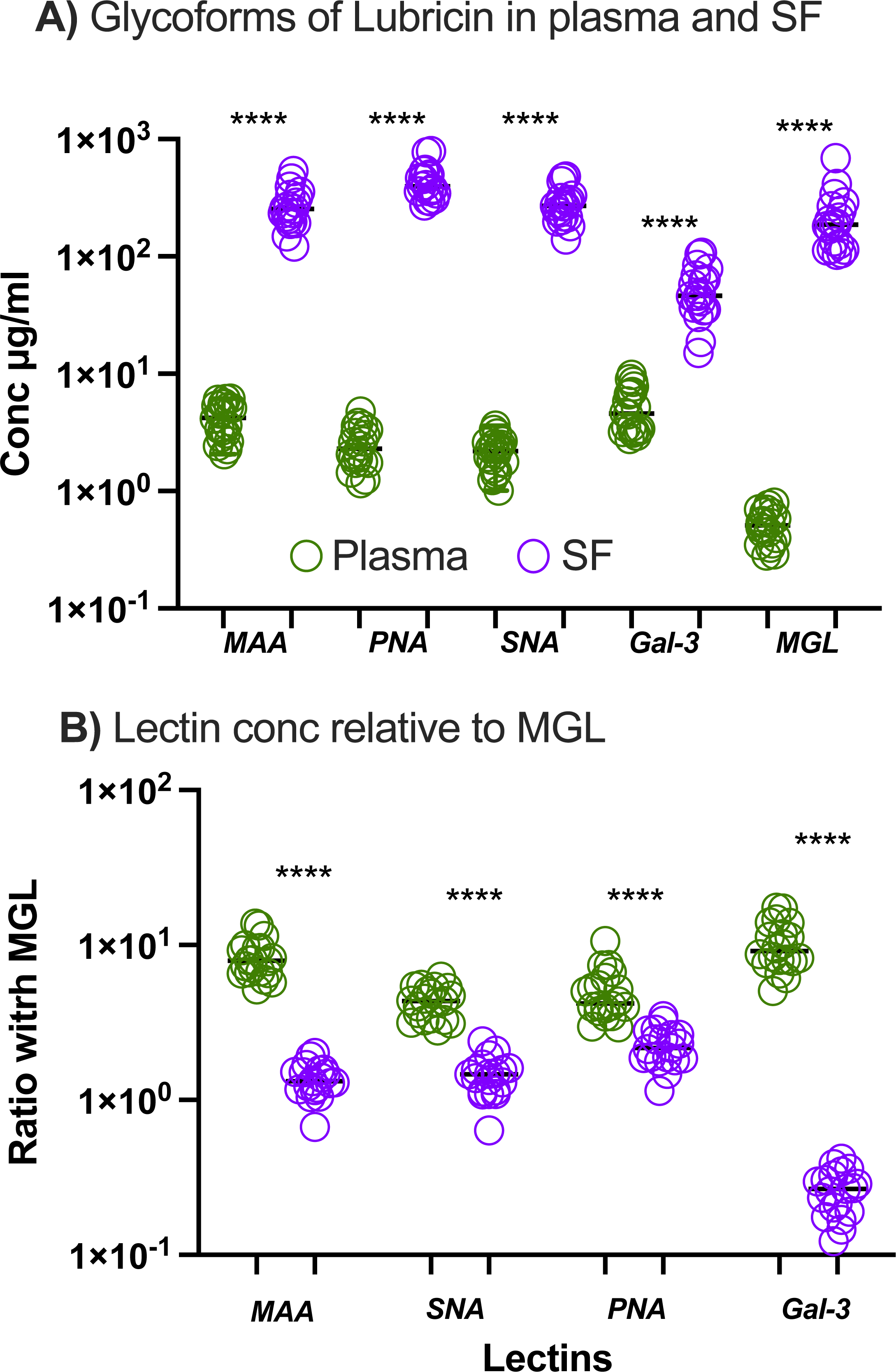
Lubricin molecular map. Schematics of lubricin with the *N*-terminal region containing the two Somatomedin-B (SMB)-like domains, STP-rich (Ser, Thr, and Pro amino acid) domain subjected to *O*-GalNAc glycosylation and the C-terminal region containing the Hemopexin (PEX)-domains. The lower image displays lubricin with its glycosylation using the SNFG cartons for monosaccharides^47^. The image is created using BioRender.com.

Lubricin is heavily glycosylated in its mucin-like domain with *O*-linked glycans^26^, that counts for > 50% of its mass^27,28^. Lubricin in its largest splice form is translated into 1404 amino acid residues, where the STP-rich domain consists of several imperfect repeats of the amino acid sequence KEPAPTTP^13,29^. The glycosylation of lubricin occurs on its STP-rich domain, that have shown to harbor more than 160 binding sites for *O-*linked glycosylation including sialylated/non sialylated core-1 *O*-glycans (Galβ1-3GalNAcα1-), and core-2 structures such as NeuAcα2-3Galβ1-3(NeuAcα2-3Galβ1-4GlcNAcβ1-6)GalNAcα1-^28,29^. In OA, there is an increase of truncated structures such as Tn-antigen (GalNAcα1-Ser/Thr) in SF lubricin^10^. Lubricin with truncated glycosylation acts as a signaling molecule that stimulates synoviocytes influencing the level of inflammatory cytokines, chemokines, and growth factors^10^.

In this report we set out to investigate the different glycoforms of lubricin found in SF^10^ and plasma, in respect to OA. In order to detect differences in glycosylation we developed and validated a Fluorescent Immuno-Lectin Assay (FILA), to ensure that we had the sensitivity required.

## Results

### Overview results

We selected five lectins including Macrophage Galactose-type lectin (MGL, GalNAcα-1, Tn-antigen), *Sambucus Nigra* Agglutinin (SNA, NeuAcα2-6, only found as NeuAcα2-6GalNAc in lubricin)^23^, *Maackia Amurensis* Agglutinin (MAA, NeuAcα2-3Galβ1-), Peanut Agglutinin (PNA,Galβ1-3GalNAcα1-, T-antigen, core 1) and Galectin-3 (Gal-3, Galβ1-4GlcNAc-, *N*-acetyllactosamine type 2), corresponding to glycoepitopes previously found on lubricin^13^, and attached them covalently to fluorescent nanoparticles. These lectin-nanoparticles were then used to measure the amount of glycoepitopes on lubricin captured by mAbs against lubricin (Figure 2A). We validated this FILA assay based on recommended tests such as spike-and-recovery, dilution-recovery, and inter-/intra-plate variation^30,31^. In addition, we benchmarked the FILA assay using SF from OA patients and compared it to a quantitative mass spectrometric Selected Reaction Monitoring (SRM) method for individual glycans from lubricin (Figure 2B-C). The validated method was then used for comparing lubricin glycosylation in plasma versus SF as well as comparing lubricin glycosylation in plasma from OA-patients and controls.

**Figure 2:**
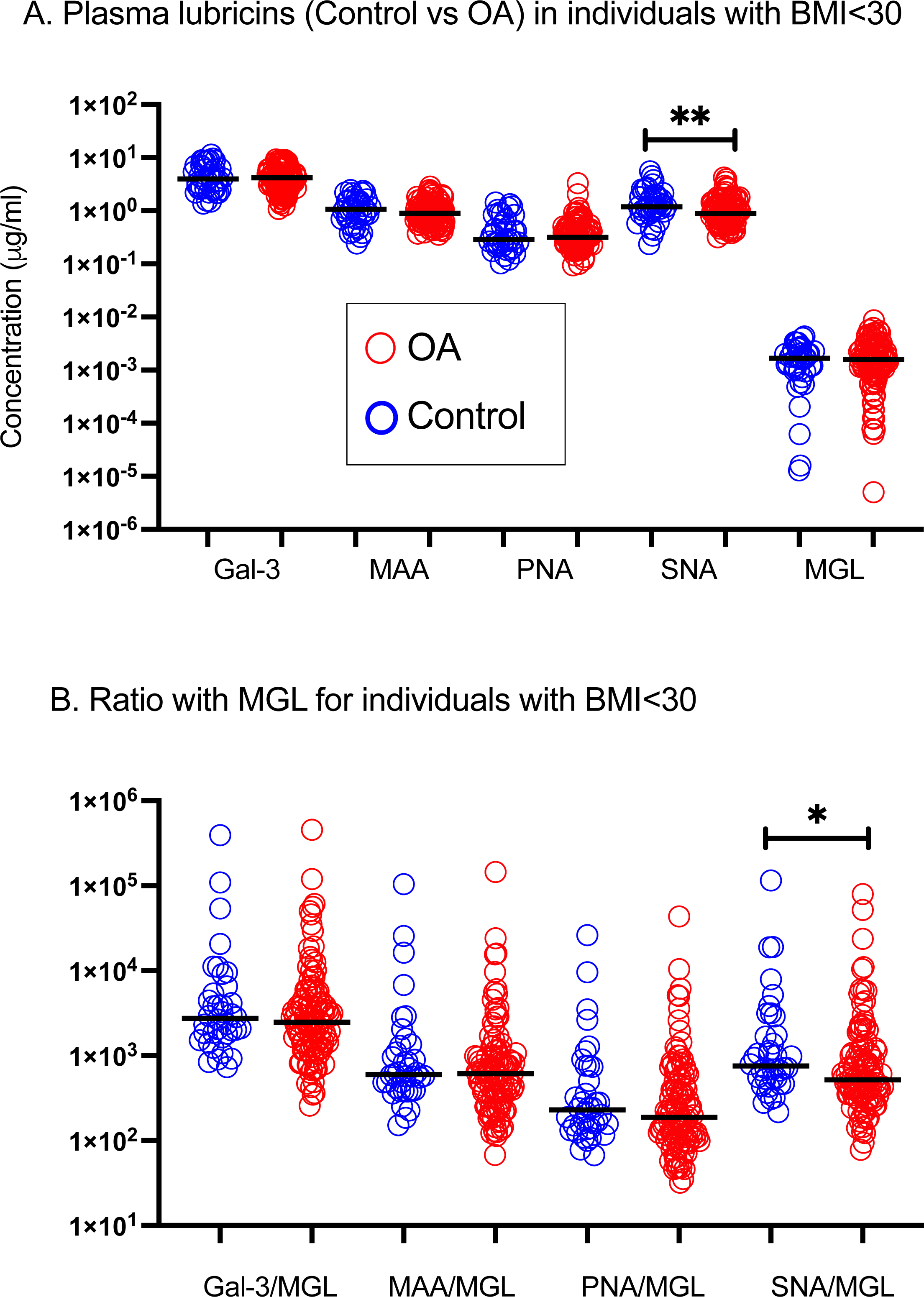
Overview of experiments. **A**. Schematic of Fluorescent Immuno Lectin Assay (upper) for measuring lubricin glycoforms sandwiched between lubricin mAb attached via biotin to streptavidin coated plates and Eu^3+^ fluorescent nanoparticles coated with lectins. The lower panel showing response from FILA by the different lectins comparing OA and Control plasma (actual data from pilot study). **B**. Example of LC-SRM results quantifying glycans present on SF lubricin from an OA patient. **C**. Correlation between FILA and LC-SRM from SF lubricin evaluated using linear regression analysis. For monosaccharide cartoon see Figure 1. The image is partly created using BioRender.com.

### Validation of FILA for screening of plasma lubricin glycosylation

For the validation of the lectin immunoassay we performed a spike-and-recovery test using a low (1x), medium (3x), and high spike (9x) (Table 1). Overall, for the range tested and for all lectins we found a good recovery (92%). In general, we found that the recovery was lower the higher the spike concentration, with an average of 117% (1x) versus 72% (9x), indicating a suppression of signal for higher concentration.

**Table 1:** Validation of FILA for detection of lubricins glycoforms in plasma using 5 different lectins.

| **Lectins** | **Sample** | **Spike and recovery (%)** |  |  | **Dilution** | **Plate CV (%)** |  |
| --- | --- | --- | --- | --- | --- | --- | --- |
|  |  | **Low**<br>**(1x)** | **Medium**<br>**(3x)** | **High**<br>**(9x)** | **Recovery (%)** | **Intra-** | **Inter-** |
| **MAA** | 1 | 120 | 121 | 85 | 100-111 | 11 | 22 |
|  | 2 | 189 | 116 | 96 | 80-100 |  |  |
| **PNA** | 1 | 95 | 77 | 58 | 100-170 | 20 | 17 |
|  | 2 | 154 | 87 | 62 | 100-117 |  |  |
| **SNA** | 1 | 70 | 106 | 81 | 95-103 | 19 | 23 |
|  | 2 | 84 | 68 | 72 | 91-119 |  |  |
| **Gal-3** | 1 | 131 | 83 | 82 | 81-113 | 10 | 14 |
|  | 2 | 145 | 86 | 86 | 73-102 |  |  |
| **MGL** | 1 | 77 | 81 | 52 | 91-157 | 10 | 21 |
|  | 2 | 105 | 58 | 47 | 71-125 |  |  |
| **Average** |  | 117 | 88 | 72 | 88-122 | 14 | 19 |

Extreme high value of recovery for 1x was found for MAA (189%), and extreme low values of recovery were found for PNA and MGL with high suppression of the 9x spike of 58% and 47%, respectively. To test the linearity of dilution, we performed an 8 times 1:2 serial dilution of plasma samples starting from 1:150 for all lectins except MGL (1:3 instead) and we found a recovery of 88% - 122%. Intra-plate reproducibility was found to be slightly lower (14% CV) compared to Inter-plate (19%) (Table 1).

### Benchmarking FILA with quantitative glyco SRM-MS

Lubricin glycans in SF are predominantly made up of simple *O*-linked glycans, consisting of Tn antigens, core 1 and core 2 (Galβ1-3[GlcNAcβ1-6]GalNAc with/without sialic acid and sulfate. The whole glycan set would comprise of a total of 15 structures, where three pairs are sequence isomers^23^. This relatively small number allows screening of the *O*-linked glycans released from lubricin to obtain quantitative information by mass spectrometry using LC separation in combination with SRM as we have shown previously^23^ (Figure 2B). We used LC-SRM to benchmark the FILA for lubricin glycoforms (Figure 2C). For this we used SF from OA patients, and isolated lubricin by ion exchange chromatography before the glycans were released. The variation of lubricin glycan structure found in OA patient’s synovial fluid was used to correlate the response between LC-SRM and the FILA. We usually found a good agreement between the two assays (Figure 2C), with a regression for all SRM/FILA above 0.6. The lowest regression was found for SNA, that also displayed the lowest variation in regards of level between the different samples, both found by SRM and FILA. The low correlation was probably a reflection of that the level of SNA-glycoform did not vary enough between our samples, superseded by the precision of the measurement.

### Lubricin glycoforms in OA patients’ plasma are different compared their synovial lubricin glycoforms

We performed the FILA using the five lectins and selected matched plasma/SF from 19 OA patients (Figure 3A). What was obvious from the analysis was that for all glycoforms, the level in SF was 1-3 order of magnitudes higher compared to plasma. For most of the glycoforms the levels in plasma were found to be below 10 μg/ml, while in SF glycoforms were usually detected at above 100 μg/ml, except for the glycoform detected by Gal-3 that was slightly lower. The highest difference in the glycoform levels between SF and plasma were found using the MGL-lectin (Tn-antigen), where the levels in plasma were less than 1 μg/ml. To appreciate differences in lubricin glycosylation between SF and plasma, we generated the ratio of all glycoforms detected with MAA, PNA, SNA and Gal-3 against glycoforms detected by MGL (Figure 3B). We found that for all epitopes, the ratio was higher in plasma compared to SF, and thus highlighting that the glycosylation of plasma derived lubricin is markedly different to SF lubricin.

**Figure 3:**
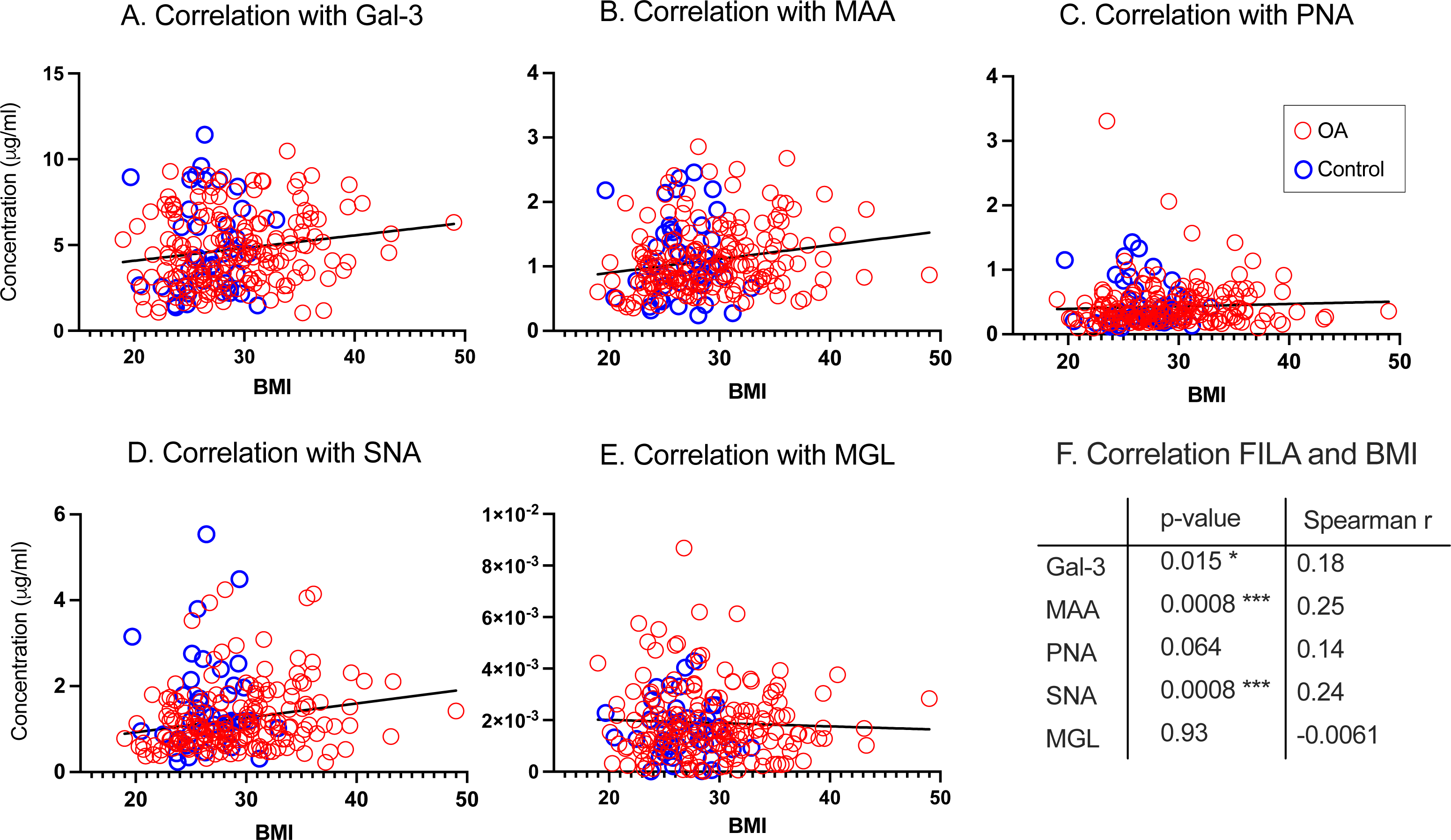
FILA results showing quantitative and glyco-qualitative differences comparing plasma and SF lubricin glycoforms from OA patients. **A.** Concentration of individual lubricin glycoforms using FILA comparing plasma lubricin and matched SF lubricin from late-stage knee OA patients (N=19). **B**. Ratios between individual lectin concentration and MGL concentrations to generate a concentration independent value to display major differences in glycosylation between SF and plasma lubricin. The p-value has been calculated with Wilcoxon matched pairs signed rank test. **** means p< 0.0001. Unmarked difference means non-significance (p>0.05).

### Plasma lubricin glycoforms in OA and control

To identify differences in lubricin glycoforms in plasma between controls and OA patients, we selected non-obese (BMI<30) OA patients due for Total Knee Replacement from two orthopedic clinics in Sweden (N=183) and controls (N=38). The selection criteria for controls were individuals with no self-reported history of recuring joint problems and not diagnosed with having previous OA. Females were overrepresented in OA patients (59%) reflecting the increased incidence of OA in females in the population. However, no significance was detected in the gender, age and BMI distribution between our OA patients and controls.

Using FILA, SNA, MAA, PNA and Gal-3, the concentration of glycoforms of lubricin was found to be around 10 μg/ml, while the MGL glycoform was found to be 1-4 order of magnitudes lower (Figure 4A). The statistic evaluation shows that plasma lubricin levels of lectin-binding glycoforms (MAA, Gal-3, PNA, and MGL), in OA and controls were not significant showing p-values ranging between 0.20 to 0.74 (Supplementary Table 3). Sialyl-Tn-antigen and related structure with 6 linked sialic acid detected by SNA lectin showed a decrease in OA (p=0.0023). We also performed the calculation of glycoform ratio dividing the individual lectin concentration using FILA with the MGL lectin. Also using this measure, we found that the ration between SNA/MGL was significantly different (p=0.0178), while the other lectin ratios were not (Figure 4B). This data suggest that OA patients display a decreased level of plasma lubricin glycoforms containing epitopes for SNA compared to controls.

**Figure 4:**
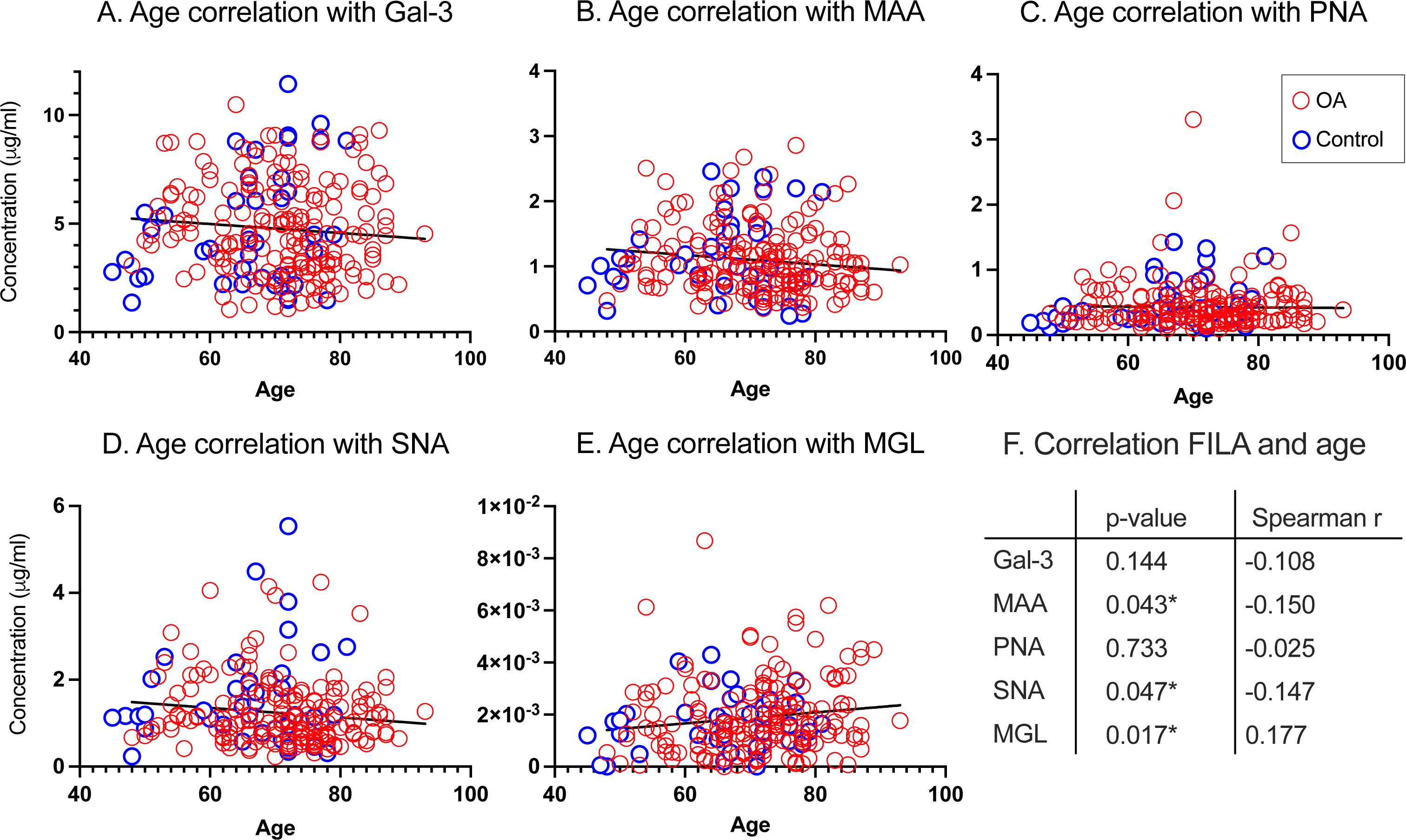
Plasma lubricin glyco-epitopes measure by FILA displaying OA patients versus controls. **A**. FILA showing concentration of lubricin glycoforms measured by various lectins. ** Marked for SNA (p=0.0023). **B**. Ratios between individual lectin concentration and MGL concentrations to generate a concentration independent value to display major differences in glycosylation between OA and control plasma lubricin. * Marked for SNA/MGL (p=0.0178). Non-parametric Mann-Whitney U-test (unpaired) was used to calculate the p-values. The data displays age, gender and BMI matched (BMI<30) late-stage OA patients (N=108) versus controls (38). Unmarked difference means non-significance (p>0.05).

### Lubricin glycoforms in plasma correlate with increased BMI and age

We also addressed if there was a correlation between lubricin glycoforms and BMI. Selecting both obese and non-obese patients and controls from the biobank, significant positive correlations between BMI and lubricin glycoforms was detected using lectins Gal-3 (p=0.016), MAA (p=0.0033), and SNA (p=0.0018), but not for MGL and PNA (Figure 5). These data indicates that BMI/overweight is associated with increased sialylation (MAA and SNA) and complexity (Type 2 *N*-acetyllactosamine elongation, Gal-3) of lubricin.

**Figure 5:**
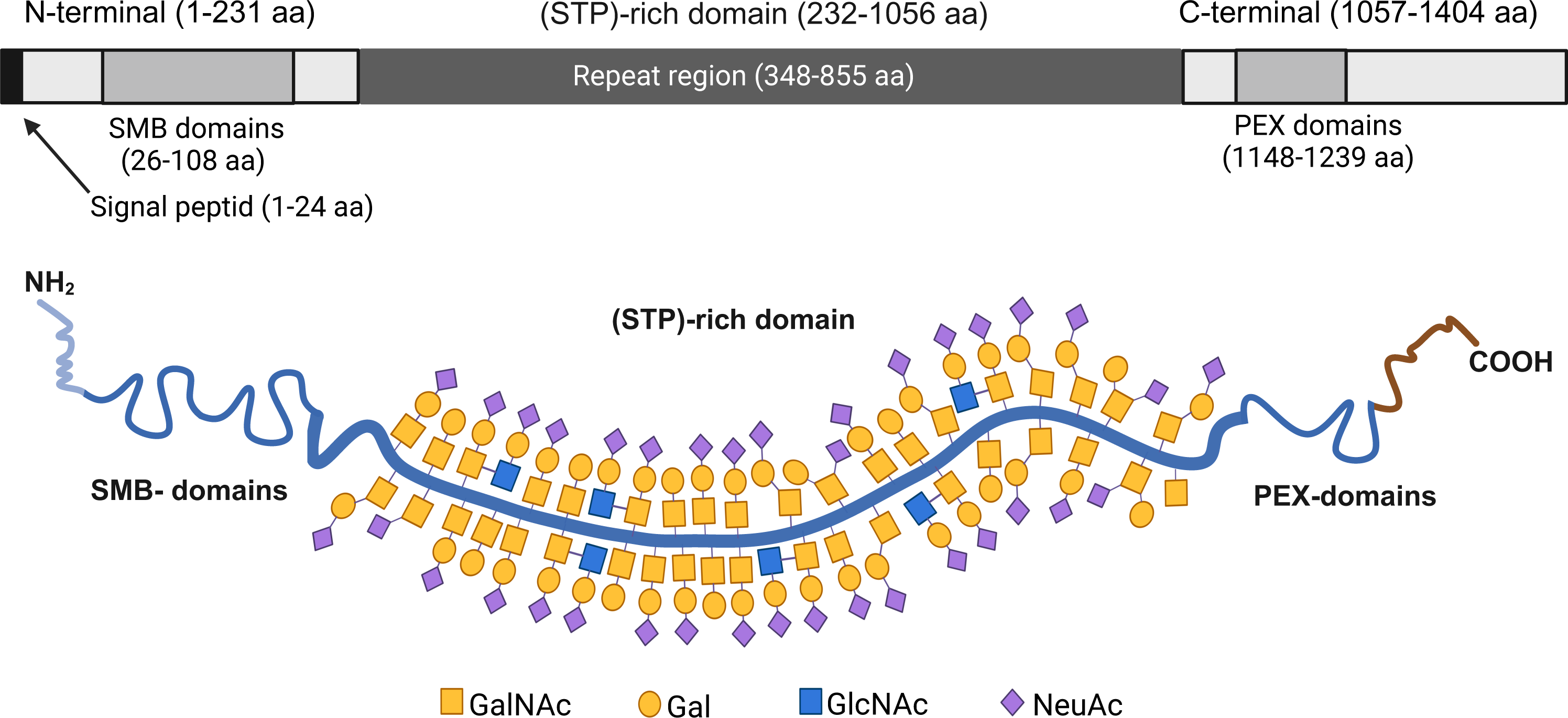
Correlation between plasma lubricin FILA glycoforms and BMI. **A-E.** The FILA results from individual lectins from plasma lubricin from late-stage knee OA ((N=183) and controls (N=41), including both obese (BMI ≥30) individuals and non-obese (BMI<30) from the biobank. Inserted is also trendlines showing negative or positive correlation between individual lectins and BMI. **F.** Correlation between lectins and BMI using Spearman r.

We also correlated lubricin glycoforms in plasma with age. Significant negative correlations between age and lubricin glycoforms in our OA patients was detected using lectins MAA (p=0.043), and SNA (p=0.0047) and positive correlation for MGL (p=0.017). Gal-3 and PNA did not show statistical correlation with age (Figure 6).

**Figure 6:**
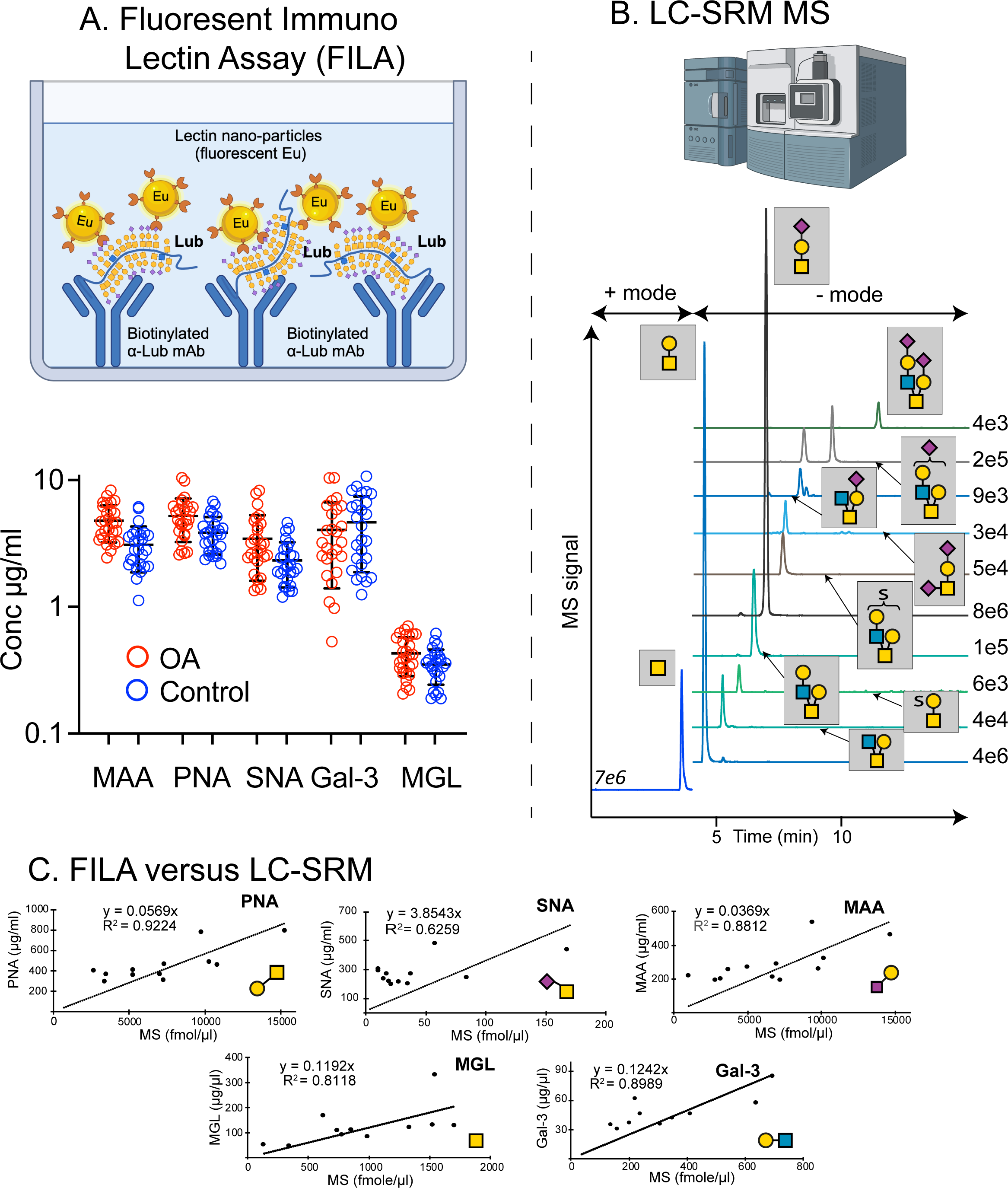
Correlation between plasma lubricin FILA glycoforms and Age. **A-E.** The FILA results from individual lectins from plasma lubricin from late-stage knee OA ((N=183) and controls (N=41). Inserted is also trendlines showing negative or positive correlation between individual lectins and Age. **F.** Correlation between lectins and BMI using Spearman r.

Interestingly, the positive impact of BMI in regards of sialylation, measured both by MAA and SNA appeared to be counteracted by age, since there was a slight negative correlation between both SNA and MAA and age.

## Discussion

We validated the lectin FILA according to selectivity, accuracy and precision and dilution linearity^32^, and cross validated the FILA with SRM of oligosaccharides from lubricin in synovial fluid using patient material. Overall and for individual lectins we could identify that we were within or close to recommendation (±20%) for spike and recovery, dilution recovery and inter-/intra-plate CV^32^. The data also showed good agreement between SRM and FILA. These validation data provided us with confidence that our FILA assay was providing a good picture of lubricin glycosylation in plasma.

Our results demonstrated that both the level and the glycosylation of lubricin from plasma and SF is different. Our FILA data suggested levels of lubricin of >100 μg/ml in SF and < 10 μg/ml in plasma. These concentrations are consistent with what have been reported in human SF and plasma previously^33-35^. The glycosylation of SF lubricin has been thoroughly characterized, demonstrating predominant level of Tn-antigen and un-sialylated/mono-sialylated core 1 glycans and lower levels of disialylated structures and core-2 glycans^13,23^, consistent with the FILA data. The FILA data from plasma suggest that lubricin has higher level of sialylation, both 3- and 6-linked relative to SF lubricin (Figure 3). The relative increased binding to PNA (core 1, T-antigen) and Gal-3 (core 2, Type-2 *N*-acetyllactosamine) is on the expense of a relative decrease of Tn-antigen in plasma compared to SF (MGL-binding) (Figure 3 B). Both core-1 and core-2 structures in plasma lubricin are then the target for the increased sialylation found in plasma compared to SF. The difference in glycosylation of lubricin between the two fluids suggest that the lubricin in plasma are mainly made up of lubricin secreted from different tissue origin(s) than SF lubricin. The cells that are synthesizing SF lubricin are expected to be found in the joint area such as synoviocytes and chondrocytes, while plasma lubricin would originate from high expressing tissue such as liver^36^ that will secrete the protein into the blood. Due to the high content of Tn-antigen on SF lubricin shown here, it would be predicted that Tn-containing lubricin escaping the joint capsule into the blood would have a short biological half-life, quickly removed by the Asialoglycoprotein receptor (ASGPR) expressed in liver^37^ . Hence, it is possible that lubricin SF glycoforms escaping the synovial areas may have different fate as they start to circulate in the plasma compared to glycoforms with less Tn antigens and higher sialylation synthesized by other cells.

The change of plasma glycosylation in OA with decreased amount of SNA epitopes is quite different compared to OA regarding SF lubricin where the glycans are shown to be more truncated and expected to have more Tn antigens compared to controls^10^. The origin of this lubricin SNA depleted glycoform found in the OA-cohort ‘s plasma is unknown and whether it is caused by alterations in catabolic and/or anabolic processes. Interestingly, the data from the comparison between SF and plasma lubricin (Figure 3B) indicates that SF lubricin has a relative lower content of SNA epitopes compared to plasma lubricin. The small change in SNA binding indicates that this alteration of lubricin in plasma could be included in a combined assay of biomarkers for OA diagnostic/prognostic, but it would not suffice by itself.

Lubricin level in plasma has been suggested to correlate with Joint Space Narrowing (JSN)^38^, but so far, a specific glycoform of lubricin has not been shown to be related to OA. Further differentiation between plasma and SF derived lubricin would be possible with additional molecular selection. It has been shown that the splice-form of lubricin produced by hepatocytes is shorter compared to the SF form, with difference in the deletion of a peptide segment in the non-glycosylated region^22,39^. Identifying splice form of lubricin found in plasma may be an additional criterion to trace the origin of lubricin in plasma. The biomedical background of the potential change of glycoforms in OA plasma, needs further investigation, whether it is originating from a local inflammation of the joint, or it is due to a more profound systemic alteration of the health status in our patients that were all from the late stage of OA.

The correlation between lubricin lectin binding in plasma and BMI, suggests that metabolic syndrome could a main driver of altered *O*-linked glycosylation of lubricin in plasma, especially the increase of sialic acid with BMI. A potential connection between lubricin and BMI is a systemic inflammation. In this case the inflammation is associated with overweight^40,41^. In other causes of cause of inflammation, such as an an acute sepsis model in mice, hepatic lubricin displays the highest upregulation of all proteins^42^. This may suggest that lubricin has an acute phase protein like behavior. The influence of *O*-linked glycosylation remodeling in hepatic acute phase glycoproteins has not been widely studied. However, remodeling of *N*-glycosylation of acute phase proteins has been reported in both chronic and acute inflammation^43^.

In regards of aging, the plasma lubricin glycosylation was indicated to display decreased sialylation (MAA and SNA) with a concomitant upregulation of the Tn antigen (MGL). Interestingly SNA (p=0,0023) and MAA (p=0.20) was the lectins that displayed the highest significance in OA versus control plasma. This indicates that aging and OA manifested similarly in regards of plasma lubricin glycoform alteration, in the opposite direction to BMI, where sialylation appeared to increase with increased BMI. The relation of *O*-linked glycosylation and age in plasma has not been widely studied. Studies on plasma N-linked glycosylation and age shows that the effect varies depending on protein (and potentially origin)^44^.

We conclude that using glyco-analytics, we have been able to trace that lubricin in plasma and SF are of different , since there are significant differences in the glycosylation. Hence, the glycosylation alteration of lubricin as we found in OA SF^10,23^would have limited impact on the glycosylation of lubricin in plasma, and we only found glycoforms detected by SNA to be altered in OA plasma. Our data also suggest that lubricin in plasma is altered by age and in metabolic syndrome manifested as increased BMI. Lubricin has been indicated to be involved in both atherosclerosis^45^ and in high-fat-diet induced glycose intolerance^46^. Changed glycosylation of lubricin in these diseases are likely to contribute to the disease progress.

## Materials and Methods

### Ethics declarations and clinical samples

Plasma samples and their controls were from a biobank maintained by Sahlgrenska University hospital (Gothenburg, Sweden). All patients and participants had given informed consent. Ethical permission for patient screening was obtained from the ethical board in Gothenburg, Sweden (ethical application 172-15). Analysis of OA biobank samples has been approved by REK (470919) and handling of personal data by NSD (431575). The study was performed using samples from 183 OA patients and 41 controls. The information about the patients is summarized in Table 2.

**Table 2:** Patient Information:

| **Patients and control BMI<30** | **Control** | **OA** |
| --- | --- | --- |
| **Age (years):** median (span) | 67 (45 – 81) | 74 (48 – 89) |
| **BMI:** median (span) | 26 (20 – 30) | 26 (19 – 30) |
| **Total:** n (% of total) | 38 (26%) | 108 (74%) |
| **Female:** n (% within group) | 19 (50%) | 64 (59%) |

### MS-FILA cross validation of lubricin glycosylation

Lubricin from OA patients (n = 20) were isolated from SF samples by anion exchange chromatography as previously described^23^. Lubricin containing fractions were concentrated using 100 kD spin filters (0.5 ml Amicon Ultra, Millipore) and salt exchange was performed with 3 x 0.5 ml 0.1M NH_4_HCO_3_, followed by speedvac to dryness. The standard mixture contained four standards: GalNAc (Tn) was from Sigma-Aldrich (St Louis MO, US), Galβ1-3GalNAc (T) from Dextra (Reading, UK), and NeuAcα2-3Galβ1-3GalNAc (ST) and Sulfo Lea (HSO_3_-3Galβ1-3(Fucα1-4)GlcNAc) were purchased from Carbosynth (Berkshire, UK). The *O*-linked oligosaccharides were released as alditols by reductive β-elimination in 100 μl sodium borohydride (1.0 M) in sodium hydroxide (0.10 M) at 50°C overnight followed by cleanup using 150 μl cation exchange media (AG50WX8, Biorad, Hercules, CA, US) on top of C18 SPE columns (Strata C18-E, 100 mg, Phenomenex, Torrance, CA, US). The oligosaccharides were dried in the speedvac, followed by repeated additions of 5 x 50 μl methanol and dried in speedvac, to evaporate borate methyl esters. Glycan standards were reduced to alditols at same conditions as above for 3 hours or overnight. The oligosaccharides were dissolved in 100 μl H_2_O, followed by injection (8 μl) on a Waters UPLC-MS/MS (Acquity Xevo TQ-S) triple quadrupole mass spectrometer. Oligosaccharides were separated on porous graphitized carbon columns (100 x 2.1 mm, 3 µm particles, Hypercarb, ThermoFisher Scientific, Waltham, MA) kept at 25°C. The gradient (36 min) consisted of 0-20 min 0-40% B (A: 10 mM ammonium bicarbonate, B: 80% acetonitrile in 10 mM ammonium bicarbonate), 20-23 min 40-100% B, 24-26 min wash with 1% HAc, then equilibration 26-36 min with 100% A. The flow rate was kept at 150 µl/min. The ESI capillary was kept on 2.5 kV. The source was at 150°C, and the cone at 40V. The samples were injected twice to cover a large range of glycans. The instrument was run in positive mode the first 4.35 min for analysis of GalNAcol (elution time 4 min). Positive mode transitions were the same for injections analyses, and covered the following six transitions at collision energy (CE)=30% and with a dwell time of 0.052 ms per transition during every cycle: *m/z* 224 to 182 ([M+H]^+^-C_2_H_2_O), *m/z* 224 to 206 ([M+H]^+^-H_2_0) and also the corresponding C13 isotope transitions: *m/z* 225 to 183, *m/z* 225 to 207, *m/z* 246 to 182 ([M+Na]^+^-C_2_H_2_O) and *m/z* 246 to 206 ([M+Na]^+^-H_2_O). During the second period of the chromatographic run the instrument was run in negative mode (4.35-20 min), covering nine transitions per run and at a dwell time of 0.032 ms: *m/z* 384.2 to 204.1 (CE=15); *m/z* 464.1 to 241.1 (CE=40); *m/z* 464.1 to 302.1 (CE=40); *m/z* 587.2 to 384.2 (CE=20); *m/z* 587.2 to 407.2 (CE=20); *m/z* 610.2 to 464.2 (CE=40) ; *m/z* 667.2 to 241.1 (CE=45); *m/z* 667.2 to 505.2 (CE=45); *m/z* 675.2 to 290.1 (CE=30); *m/z* 749.3 to 569.2 (CE=30); *m/z* 755.2 to 465.2 (CE=50); 829.2 to 667.2 (CE=50); *m/z* 878.3 to 290.1 (CE=40); *m/z* 966.3 to 290.1 (CE=50);*m/z* 1040.4 to 290.1 (CE=45) and *m/z* 1331.5 to 290.1 (CE=60). Collision energies were optimized using syringe infusions of saliva mucin oligosaccharides, fetuin and porcine gastric mucin oligosaccharides. A dilution series (1:2) of the standard mixture was made (0.10-90 pmol on column), with analyses of 1-2 standards between every three samples. Standards were analyzed using the same method as for samples, although 2 μl were injected.

### Biotinylation of antibodies

Sulfo N-hydroxysuccinimide Biotin (sulfo-NHS-biotin) (Thermos Fisher, Waltham, Massachusetts, US) was used for the biotinylating of antibodies. In-house mouse IgG antibodies 1E12 (0.42 mg, for plasma-SF comparison) and 14G10 (0.71 mg, for plasma OA-Control comparison) against mucin domain of lubricin was biotinylated in 2.5 ml of PBS with 20-fold molar excess of sulfo-NHS-biotin compared to antibody. The biotinylation reaction was allowed to proceed for two hours. Excess biotin was removed using a PD-10 column (Cytiva, Little Chalfont, UK) according to manufacturer’s recommendation.

### Lectin coating of Eu^3+^ nanoparticles

Lectins (Vector labs, Newark, CA, US) were coated covalently attached to 97 nm Fluoro-Max Dyed Carboxylate-modified Microparticles (Thermo Fisher) (20 µl). The particles were applied to 1 Nanosep 300kDa Omega (Thermo Fisher) centrifugal filter. The Nanosep tube was centrifuged until dry at 8000 rpm for five minutes. Filter was washed twice with 200 µl of conjugation buffer (2-(N-morpholino)ethanesulfonic acid (MES) buffer, 50 mM, pH 6.0) and finally resuspended adding 150 µl of conjugation buffer followed by sonication with 40 pulses using an ultrasonic probe. Particles was activated adding sulfo-NHS (230 mM dissolved in conjugation buffer, 7.4 µl) and 1-ethyl-3-(3’dimethylaminopropyl)-carbodiimide (Sigma-Aldrich, 52 mM dissolved in conjugation buffer 10.4 µl) and agitated for 15 min at room-temperature using vortex. Lectins (0.5 mg/ml dissolved in PBS, 75 µl) were added to 50 µl of particle solution and the volume was adjusted to a total of 156 µl using 500 mM MES (4.5 µl), 3.5 µl 3.0 M NaCl, 17 µl water and 6.5 µl carbonate buffer 0. 50 M, pH 9.8 and incubated and repeatedly vortexed for another 30 min. The solution was stored over night at +4 °C in rotary mixer with the exception for SNA that was just stored in +4 °C without mixing. The reaction mixture was removed on the next day using one NanoSep 300 and centrifugated (8000 rpm) for five min followed by washing with storage buffer twice (100 μl) (25 mM Tris, 150 mM NaCl, 0.1% NaN_3_ pH 7.8) and centrifugation at 8000 rpm for five min. Storage buffer (100 µl) was added on the filter and the lectin coated particles was resuspend by ultra-sonication.

### The fluorescent lectin-immune assay (FILA)

Streptavidin coated micro-plates (SA96-plates, Kaivogen, Turku, Finland) were prewashed 1x with washing buffer (Kaivogen, Turku, Finland). Biotinylated mAb 1E12 and 14G10 targeting lubricin was added to each well (25 ng/well, 25 μl) and plates were incubated for one hour in RT. After incubation the wells were washed 2x by washing buffer and plasma samples were added in triplicates. Each sample was diluted (1:150 and 1:400 or 1:5 and 1:10 for MGL) in red buffer. In addition, a serial dilution (1.3×10^-1^ µg/ml up to 2.5×10^-4^ µg/ml) of recombinant lubricin-(rhPRG4) stock conc.1300 µg/ml) were added to the plate in triplicates. Plates were then incubated for one hour at room temperature. After the incubation all plates were washed 2x and lectin Eu^3+^-nanoparticles (1×10^7^ particle/well, 25 µl). The plates were incubated for one hour at RT and washed 6x with washing buffer. The fluorescence was read using VictorNivo multimode plate reader (PerkinElmer). The measurement operation was time-resolved fluorescence (TRF) with excitation 320 nm, and emission 615 nm wavelength. The concentration of lubricin glycoforms was calculated using MyAssays analysis software (MyAssays Ltd, Brighton, UK).

### Validation of immune assay

For the validation of immune assay both spike and recovery test and test for the linearity of dilutions were performed. In addition, a system-control was used for calculation of intra- and inter-assay coefficient of variations (CVs) according to recommendation^31^.

For spike and recovery test, diluted plasma was used adding of known amounts of rhPRG4 (spike) to selected samples. Plasma diluted was spiked with low (0.55 µg/ml), medium (1.7 µg/ml), and high (5.0 µg/ml) concentrations using rhPRG4. For recovery 1:150, 1:300, 1:600, 1:1200 diluted plasma was used and added to the plate in triplicates. In case of MGL selected plasma samples were spiked with low (0.3 µg/ml), medium (1.0 µg/ml), and high (2.9 µg/ml) concentrations using rhPRG4 and for dilution recovery 1:3, 1:6, 1:12, 1:24 diluted plasma were used. Intra-plate CV% was determined for identical samples in triplicates from the same plate. Inter-plate CV% was determined from same system-control sample in different plates.

### Statistics

In total 224 samples were used for analyzing plasma lubricin glycoforms and to determine the differences in the subgroups: OA (n=183) and controls (n=41). Statistical analyses were performed using statistical software: SPSS (IBM, Armonk, NY, US) and GraphPad Prism 9 (Dotmatics, CA, US). Continuous variables were summarized as mean and standard deviation or median and minimum-maximum. Assumptions such as outliers and normal distribution (Shapiro-wilk test) were considered, where outliers were kept. All data were normalized and non-parametric Mann-Whitney U-test (unpaired) or Wilcoxon matched pairs were used to determine if there were differences in glycoforms of lubricin between subgroups (for each glycoform separately). The significant level was considered as p<0.05 (two-tails).

Quantitative analysis of samples and standard curves were carried out regarding different types of lectins. For the calculation of MGL concentration in samples linear regression was used and for other lectins four parameter logistic (4PL).

## Supplementary material

**Supplementary Data 1**: Raw data from FILA for all individuals

**Supplementary Data 2:** Raw data from FILA for individuals BMI<30

**Supplementary Tables: Table 1:** Comparison of FILA levels between OA and control plasma lubricin glycoforms in individuals with BMI<30 and for all individuals. Table 2: Correlation between lubricin glycoforms and BMI. Table 3: Correlation between lubricin glycoforms and age.

## Data availability statement

Analytical data about individual patient FILA results are included in the Supplementary Material. Inquiries about additional patient meta data can be directed to the corresponding author.

## Author Contributions

ARA and NGK planned, and ARA performed the validation of the FILA. ARA planned and executed the OA and control FILA experiments and performed the statistical evaluation of the results. JH, KG and KP adopted the FILA for lubricin glycosylation analysis and JH performed the plasma-SF comparison. KT and HR performed SRM experiments. OR, LIB and TE were responsible for providing control and patient sample material. TS and GJ contributed to the design and provision of rhPRG4 for the analysis and antibody generation. ARA wrote the initial draft, and NK completed the writing with input from all contributors. NGK designed the research plan, coordinated, and directed the research, and performed the overall statistical evaluation. NK and GJ generated funding for the research. All authors critically reviewed and approved the submitted version.

## Acknowledgments

Sofia Grindberg and Paula Therese Kelly Petterssons at Danderyd’s Hospital and Lotta Falkendahl at the University of Gothenburg are acknowledged for their assistance in collecting samples. Dr Shan Huang is acknowledged in guidance in validation of the FILA. Lubris BioPharma is acknowledged for the generous gift of rhPRG4. We also gratefully acknowledge financial contribution to NGK from the Swedish state under the agreement between the Swedish government and the county council, the ALF-agreement (ALFGBG-722391), the Swedish Research Council (621-2013-5895), Petrus and Augusta Hedlund’s foundation (M-2016-0353), and AA insurance research fund (dnr 150150). GJ acknowledge funding from the National Institute of Health R01AR067748. ARA gratefully acknowledge OsloMet for providing a stipend to perform this project. Sponsors had no role in the study design, collection, analysis, and interpretation of data, nor were they involved in the writing of the manuscript and the decision to submit it for publication.

## References

1. Flowers, S. A. et al. Lubricin binds cartilage proteins, cartilage oligomeric matrix protein, fibronectin and collagen II at the cartilage surface. Sci Rep 7, 13149, doi:10.1038/s41598-017-13558-y (2017).

2. Mueller, M. B. & Tuan, R. S. Anabolic/Catabolic balance in pathogenesis of osteoarthritis: identifying molecular targets. PM R 3, S3–11, doi:10.1016/j.pmrj.2011.05.009 (2011).

3. Pereira, D., Ramos, E. & Branco, J Osteoarthritis. Acta Med Port 28, 99–106, doi:10.20344/amp.5477 (2015).

4. Tanna, S. Osteoarthritis “Opportunities to address pharmaceutical gaps”. Priority Medicines for Europe and World: a public health approach to innovation 6, 3–23 doi: 10.1186/2052-3211-8-S1-K4 (2004).

5. Khoshgoftar, M., Torzilli, P. A. & Maher, S. A. Influence of the pericellular and extracellular matrix structural properties on chondrocyte mechanics. J Orthop Res 36, 721–729, doi:10.1002/jor.23774 (2018).

6. Alexopoulos, L. G., Setton, L. A. & Guilak, F. The biomechanical role of the chondrocyte pericellular matrix in articular cartilage. Acta Biomater 1, 317–325, doi:10.1016/j.actbio.2005.02.001 (2005).

7. Alexopoulos, L. G., Williams, G. M., Upton, M. L., Setton, L. A. & Guilak, F. Osteoarthritic changes in the biphasic mechanical properties of the chondrocyte pericellular matrix in articular cartilage. J Biomech 38, 509–517, doi:10.1016/j.jbiomech.2004.04.012 (2005).

8. Wilusz, R. E., Zauscher, S. & Guilak, F. Micromechanical mapping of early osteoarthritic changes in the pericellular matrix of human articular cartilage. Osteoarthr Cartil 21, 1895–1903, doi:10.1016/j.joca.2013.08.026 (2013).

9. Mead, T. J., Bhutada, S., Martin, D. R. & Apte, S. S. Proteolysis: a key post-translational modification regulating proteoglycans. Am J Physiol Cell Physiol 323, C651–C665, doi:10.1152/ajpcell.00215.2022 (2022).

10. Huang, S. et al. Truncated lubricin glycans in osteoarthritis stimulate the synoviocyte secretion of VEGFA, IL-8, and MIP-1alpha: Interplay between O-linked glycosylation and inflammatory cytokines. Front Mol Biosci 9, 942406, doi:10.3389/fmolb.2022.942406 (2022).

11. Krasnokutsky, S., Samuels, J. & Abramson, S. B. Osteoarthritis in 2007. Bull NYU Hosp Jt Dis 65, 222–228 (2007).

12. Xia, B. et al. Osteoarthritis pathogenesis: a review of molecular mechanisms. Calcif Tissue Int 95, 495–505, doi:10.1007/s00223-014-9917-9 (2014).

13. Estrella, R. P., Whitelock, J. M., Packer, N. H. & Karlsson, N. G. The glycosylation of human synovial lubricin: implications for its role in inflammation. Biochem J 429, 359–367, doi:10.1042/BJ20100360 (2010).

14. Wang, M., Liu, C., Thormann, E. & Dėdinaitė, A. Hyaluronan and Phospholipid Association in Biolubrication. Biomacromolecules 14, 4198–4206, doi:10.1021/bm400947v (2013).

15. Atarod, M., Ludwig, T. E., Frank, C. B., Schmidt, T. A. & Shrive, N. G. Cartilage boundary lubrication of ovine synovial fluid following anterior cruciate ligament transection: a longitudinal study. Osteoarthr Cartil 23, 640–647, doi:10.1016/j.joca.2014.12.017 (2015).

16. Roughley, P. J. & Mort, J. S. The role of aggrecan in normal and osteoarthritic cartilage. J Exp Orthop 1, 8, doi:10.1186/s40634-014-0008-7 (2014).

17. Kosinska, M. K. et al. A lipidomic study of phospholipid classes and species in human synovial fluid. Arthritis Rheum 65, 2323–2333, doi:10.1002/art.38053 (2013).

18. Iqbal, S. M. et al. Lubricin/Proteoglycan 4 binds to and regulates the activity of Toll-Like Receptors In Vitro. Sci Rep 6, 18910, doi:10.1038/srep18910 (2016).

19. Jones, A. & Flannery, C. Bioregulation of lubricin expression by growth factors and cytokines. Eur Cell Mater 13, 40–45 (2007).

20. DuRaine, G. et al. Regulation of the friction coefficient of articular cartilage by TGF-β1 and IL-1β. J Orthop Res 27, 249–256 (2009).

21. Rees, S. G. et al. Immunolocalisation and expression of proteoglycan 4 (cartilage superficial zone proteoglycan) in tendon. Matrix Biol 21, 593–602, doi:10.1016/s0945-053x(02)00056-2 (2002).

22. Ikegawa, S., Sano, M., Koshizuka, Y. & Nakamura, Y. Isolation, characterization and mapping of the mouse and human PRG4 (proteoglycan 4) genes. Cytogenet Cell Genet 90, 291–297, doi:10.1159/000056791 (2000).

23. Flowers, S. A. et al. Decrease of core 2 O-glycans on synovial lubricin in osteoarthritis reduces galectin-3 mediated crosslinking. J Biol Chem 295, 16023–16036, doi:10.1074/jbc.RA120.012882 (2020).

24. Menon, N. G. et al. Proteoglycan 4 (PRG4) expression and function in dry eye associated inflammation. Exp Eye Res 208, 108628, doi:10.1016/j.exer.2021.108628 (2021).

25. Richendrfer, H. & Jay, G. D. Lubricin as a Therapeutic and Potential Biomarker in Sepsis. Crit Care Clin 36, 55–67, doi:10.1016/j.ccc.2019.08.005 (2020).

26. Rhee, D. K. et al. Consequences of disease-causing mutations on lubricin protein synthesis, secretion, and post-translational processing. J Biol Chem 280, 31325–31332, doi:10.1074/jbc.M505401200 (2005).

27. Schmidt, T. A., Plaas, A. H. & Sandy, J. D. Disulfide-bonded multimers of proteoglycan 4 PRG4 are present in normal synovial fluids. Biochim Biophys Acta 1790, 375–384, doi:10.1016/j.bbagen.2009.03.016 (2009).

28. Jay, G. D. & Waller, K. A. The biology of lubricin: near frictionless joint motion. Matrix Biol 39, 17–24, doi:10.1016/j.matbio.2014.08.008 (2014).

29. Ali, L. et al. The O-glycomap of lubricin, a novel mucin responsible for joint lubrication, identified by site-specific glycopeptide analysis. Mol Cell Proteomics 13, 3396–3409, doi:10.1074/mcp.M114.040865 (2014).

30. Brindle, E. et al. A multicenter analytical performance evaluation of a multiplexed immunoarray for the simultaneous measurement of biomarkers of micronutrient deficiency, inflammation and malarial antigenemia. PLoS One 16, e0259509, doi:10.1371/journal.pone.0259509 (2021).

31. Cox, K. L. et al. Immunoassay Methods in Assay Guidance Manual (eds S. Markossian et al.) 1-, Eli Lilly & Company and the National Center for Advancing Translational Sciences, 2004).

32. European Medicine Agency. ICH guideline M10 on bioanalytical method validation and study sample analysis. 1–45 (2022).

33. Elsaid, K. A. et al. Decreased lubricin concentrations and markers of joint inflammation in the synovial fluid of patients with anterior cruciate ligament injury. Arthritis Rheum 58, 1707–1715, doi:10.1002/art.23495 (2008).

34. Ludwig, T. E., McAllister, J. R., Lun, V., Wiley, J. P. & Schmidt, T. A. Diminished cartilage-lubricating ability of human osteoarthritic synovial fluid deficient in proteoglycan 4: Restoration through proteoglycan 4 supplementation. Arthritis Rheum 64, 3963–3971, doi:10.1002/art.34674 (2012).

35. Asfari, A. et al. Plasma proteoglycan 4: a novel biomarker for acute lung injury after pediatric cardiac surgery. Transl Pediatr 12, 1668–1675, doi:10.21037/tp-23-194 (2023).

36. Marcelino, J. et al. CACP, encoding a secreted proteoglycan, is mutated in camptodactyly-arthropathy-coxa vara-pericarditis syndrome. Nat Genet 23, 319–322, doi:10.1038/15496 (1999).

37. Morell, A. G., Irvine, R. A., Sternlieb, I., Scheinberg, I. H. & Ashwell, G. Physical and chemical studies on ceruloplasmin. V. Metabolic studies on sialic acid-free ceruloplasmin in vivo. J Biol Chem 243, 155–159 doi:10.1016/S0021-9258(18)99337-3 (1968).

38. Ritter, S. Y. et al. Mass spectrometry assays of plasma biomarkers to predict radiographic progression of knee osteoarthritis. Arthritis Res Ther 16, 456, doi:10.1186/s13075-014-0456-6 (2014).

39. Jay, G. D., Tantravahi, U., Britt, D. E., Barrach, H. J. & Cha, C. J. Homology of lubricin and superficial zone protein (SZP): products of megakaryocyte stimulating factor (MSF) gene expression by human synovial fibroblasts and articular chondrocytes localized to chromosome 1q25. J Orthop Res 19, 677–687, doi:10.1016/S0736-0266(00)00040-1 (2001).

40. Kozakowski, J., Dudek, P. & Zgliczynski, W. Obesity in rheumatological practice. Reumatologia 61, 318–325, doi:10.5114/reum/170401 (2023).

41. McCarthy, C. et al. The role and importance of glycosylation of acute phase proteins with focus on alpha-1 antitrypsin in acute and chronic inflammatory conditions. J Proteome Res 13, 3131–3143, doi:10.1021/pr500146y (2014).

42. Toledo, A. G. et al. Proteomic atlas of organ vasculopathies triggered by Staphylococcus aureus sepsis. Nat Commun 10, 4656, doi:10.1038/s41467-019-12672-x (2019).

43. Caval, T. et al. Glycoproteoform Profiles of Individual Patients’ Plasma Alpha-1-Antichymotrypsin are Unique and Extensively Remodeled Following a Septic Episode. Front Immunol 11, 608466, doi:10.3389/fimmu.2020.608466 (2020).

44. Paton, B., Suarez, M., Herrero, P. & Canela, N. Glycosylation Biomarkers Associated with Age-Related Diseases and Current Methods for Glycan Analysis. Int J Mol Sci 22, doi:10.3390/ijms22115788 (2021).

45. Seime, T. et al. Proteoglycan 4 Modulates Osteogenic Smooth Muscle Cell Differentiation during Vascular Remodeling and Intimal Calcification. Cells 10, doi:10.3390/cells10061276 (2021).

46. Nahon, J. E. et al. Proteoglycan 4 deficiency protects against glucose intolerance and fatty liver disease in diet-induced obese mice. Biochim Biophys Acta Mol Basis Dis 1865, 494–501, doi:10.1016/j.bbadis.2018.11.009 (2019).

47. Neelamegham, S. et al. Updates to the Symbol Nomenclature for Glycans guidelines. Glycobiology 29, 620–624, doi:10.1093/glycob/cwz045 (2019).

