## Supplementary tables for "A Novel Fluorescent Immuno-Lectin Assay to Identify Osteoarthritis Associated Glycoforms of Lubricin"

**Supplementary Table 1: Comparison of FILA levels between OA and control plasma lubricin glycoforms in individuals with BMI<30 and for all individuals**

| | [a]All individuals | | [b]BMI<30 | |
|---|---|---|---|---|
| **Glycoforms** | **Median** (Control / OA) | **p-value** | **Median** (Control / OA) | **p-value** |
| **Gal-3** | 4.195 / 4.519 | 0.3037 | 3.991 / 4.167 | 0.7441 |
| **MAA** | 1.007 / 1.010 | 0.9360 | 1.071 / 0.8970 | 0.2007 |
| **PNA** | 0.2998 / 0.3345 | 0.9912 | 0.2891 / 0.3179 | 0.6017 |
| **SNA** | 1.191 / 1.053 | 0.0610 | 1.198 / 0.8950 | 0.0023** |
| **MGL** | 0.00160 / 0.00158 | 0.4436 | 0.001681 / 0.001604 | 0.5524 |
| **Gal-3/MGL** | 2820 / 2567 | 0.6174 | 2744 / 2488 | 0.4276 |
| **MAA/MGL** | 610.0 / 635.0 | 0.7379 | 599.2 / 615.2 | 0.3703 |
| **PNA/MGL** | 240.4 / 217.1 | 0.3009 | 229.3 / 189.0 | 0.2007 |
| **SNA/MGL** | 793.2 / 618.6 | 0.0931 | 756.0 / 517.2 | 0.0178* |

[a] (total n=224), (Controls: n=41), (OA: n=183)
[b] (total n=146), (Controls: n=38), (OA n=108)

**Supplementary Table 2: Correlation between glycoforms and BMI**

| Correlation between **BMI** and lectins in control and OA samples | | | | | | |
|---|---|---|---|---|---|---|
| | | **Gal-3** | **MAA** | **PNA** | **SNA** | **MGL** |
| **OA** (n=183) | Spearman r | 0.1794 | 0.2453 | 0.1373 | 0.2449 | -0.00615 |
| | p-value | 0.0151 * | 0.0008 *** | 0.0638 | 0.0008 *** | 0.9342 |
| **Controls** (n=41) | Spearman r | 0.1235 | 0.04906 | 0.09891 | 0.1522 | -0.06903 |
| | p-value | 0.4418 | 0.7607 | 0.5384 | 0.3420 | 0.6681 |
| **All** (n=224) | Spearman r | 0.1653 | 0.2039 | 0.1255 | 0.1963 | -0.01051 |
| | p-value | 0.0133 * | 0.0022 ** | 0.0607 | 0.0032 ** | 0.8757 |

**Supplementary Table 3: Correlation between glycoforms and age**

| Correlation between **Age** and lectins in control and OA samples | | | | | | |
|---|---|---|---|---|---|---|
| | | **Gal-3** | **MAA** | **PNA** | **SNA** | **MGL** |
| **OA** (n = 183) | Spearman r | -0.1084 | -0,1500 | -0,02542 | -0,1470 | 0,1769 |
| | P-value | 0.1441 | 0,0426* | 0,7327 | 0,0471* | 0,0166* |
| **Controls** (n = 41) | Spearman r | 0.2459 | -0.05855 | 0.2807 | 0.01311 | 0.06493 |
| | P-value | 0.1212 | 0.7161 | 0.0755 | 0.9352 | 0.6867 |
| **All** (n = 224) | Spearman r | -0.03545 | -0.01338 | 0.02981 | -0.0149 | 0.1683 |
| | P-value | 0.5976 | 0.0455* | 0.6572 | 0.0258* | 0.0117* |
